# Priority topics for preconception care in general practice: a consensus study

**DOI:** 10.64898/2026.03.20.26348893

**Authors:** Danielle Schoenaker, Elizabeth M Lovegrove, Miriam Santer, Karen Matvienko-Sikar, Helen Carr, Nisreen A Alwan, Laura Kubelabo, Nathan Davies, Keith M Godfrey

**Author notes:** **Correspondence to:** Danielle Schoenaker, University of Southampton, MRC Lifecourse Epidemiology Centre, Tremona Road, Southampton SO16 6YD, UK.

## Abstract

**Background:** Primary care practitioners are well-positioned to support people of reproductive age in preparing for pregnancy and parenthood. Such “preconception care” is ideally delivered opportunistically during routine consultations, although limited time presents a barrier.

**Aim:** To achieve consensus on priority topics for opportunistic preconception care in general practice.

**Design and setting:** A three-step consensus study involving UK-based primary care practitioners and people of reproductive age.

**Method:** The consensus process involved: 1) identifying potential topics through literature and guideline reviews, workshops with people of reproductive age (n=15), and interviews with primary care practitioners who work in general practice (n=14); 2) prioritising topics using a Delphi survey (n=85 participants completing round one, n=63 completing all three rounds); and 3) agreeing on priority topics during an online consensus workshop (n=21 participants). Participants were recruited through a Public Advisory Group, charities, and professional organisations.

**Results:** Reviews and workshops/interviews with people of reproductive age and practitioners identified 37 potential topics. The Delphi survey and consensus workshop identified 16 priority topics. These were combined into four overarching topic areas for discussion during relevant consultations:

1. Patient knowledge of preconception health and pregnancy
2. Ideas, concerns and expectations (e.g. pregnancy intention, prior pregnancy experiences)
3. Health conditions (e.g. medication use, mental/physical health, immunisation)
4. Health behaviours (e.g. folic acid supplement use, smoking, alcohol consumption).

**Conclusion:** The agreed priority topic areas offer a structured foundation for delivering patient-centred, opportunistic preconception care in primary care. The findings support future co-development of practical tools and resources to enable routine implementation.

**How this fits in:** Preconception care improves pregnancy outcomes, but in UK general practice it is inconsistently delivered, partly due to limited time and guidance that offers little prioritisation for opportunistic consultations. This study identifies four overarching topic areas for preconception care, based on consensus among people of reproductive age and primary care practitioners. The resulting priority list offers clinicians a practical, flexible way to initiate patient-centred preconception care discussions within routine consultations.

## Introduction

Preconception health has a well-established influence on fertility, pregnancy outcomes, and the long-term health of parents and children.^1–4^ Important factors associated with adverse outcomes include for example poorly controlled chronic conditions, use of teratogenic medications, smoking and domestic abuse.^5–7^ These factors are best identified and optimised before pregnancy.^8, 9^ Primary care-based interventions have been shown to improve preconception risk factors and may reduce adverse pregnancy outcomes, including pregnancy loss and pre-eclampsia.^10^ As most people of reproductive age in high-income countries have regular contact with general practice,^11–14^ professionals in this setting are well placed to initiate opportunistic discussions about pregnancy intentions, screen for key risk factors, and offer relevant advice to support preparation for pregnancy and parenthood.

In the UK, however, preconception care is inconsistently delivered.^11, 15, 16^ Primary care professionals face competing demands, limited consultation time, and uncertainty about which issues to prioritise.^17^ Existing National Institute for Health and Care Excellence (NICE) guidance on preconception advice and management is extensive, covering more than 30 individual risk factors, but provides limited direction on how to prioritise topics in brief, opportunistic encounters.^18^ There is also limited evidence on how people of reproductive age value different aspects of preconception health,^19–21^ or how their priorities align with those of primary care professionals.^17, 22^ Without prioritisation, opportunistic preconception care risks being too narrow (e.g. focussed on only one topic), or too broad to be feasible in routine care.

To support provision of preconception care, primary care practitioners need realistic guidance that enables brief, patient-centred discussions about preparation for pregnancy and parenthood. This guidance should focus on a feasible and acceptable number of key topics and reflect both clinical priorities and the perspectives of people of reproductive age. The aim of this study was therefore to agree a set of priority topics to guide opportunistic preconception care in UK general practice, using a consensus process involving people of reproductive age and primary care practitioners.

## Methods

We conducted a three-step consensus study among people of reproductive age (defined here as 16-49 years) and primary care practitioners who work in general practice in the UK (including GPs, nurses, practice managers, among others) (Figure 1). Detailed methodology is reported in the study protocol.^23^ No protocol deviations occurred, and findings are reported in line with the ACCORD guideline.^24^

**Figure 1.**
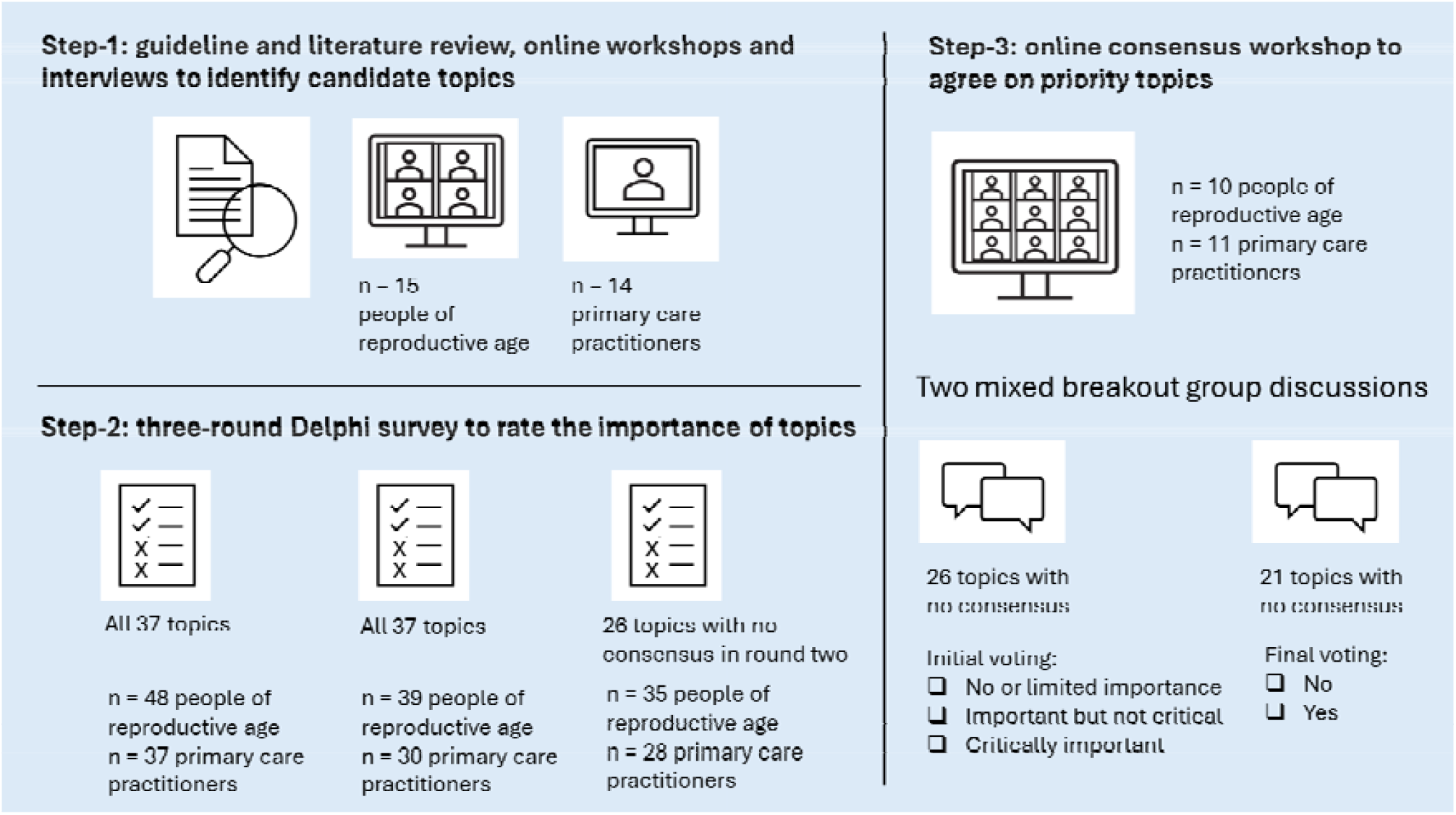
Outline of three-step study design.

### Step-1: Clinical guideline and literature review, workshops and interviews

Potential topics were identified through searches of the NICE Clinical Knowledge Summary on preconception advice and management,^18^ international preconception care guidelines,^25^ and relevant systematic reviews published in the past five years (searched October 2023; Supplementary Table 1). Identified topics were then discussed in online 1.5-hour workshops with people of reproductive age (October-November 2023) and online 30-minute interviews with primary care practitioners (November 2023-January 2024) to ensure relevance and completeness.

People of reproductive age were recruited through the study’s Public Advisory Group, who could themselves take part and invite others through their personal and community networks. Primary care practitioners were recruited through newsletter and social media posts by professional organisations. Participants were asked to provide feedback on the relevance and clarity of the topics and any missing topics. They were also asked to describe how they would decide on the importance of topics to inform prioritisation principles used in the Delphi survey. Workshops and interviews were recorded for notetaking to put findings into context.

### Step-2: Delphi survey

An online three-round Delphi survey was conducted September 2024-February 2025 among people of reproductive age and primary care practitioners. People of reproductive age were recruited from Step-1 workshop participants, further sharing of the study details by the Public Advisory Group through personal and community networks, and through national charities. Primary care practitioners were recruited from Step-1 interview participants and professional organisations (see Acknowledgements).

Participants were asked to read a hypothetical scenario and rate the proposed topics regarding importance for primary care professionals to consider each topic in the scenario, while considering prioritisation principles (Box 1). Topics were rated on a nine-point scale: ‘No/limited importance’ (scores 1-3), ‘Important but not critical’ (scores 4-6) and ‘Critically important’ (scores 7-9). In round one, participants could suggest up to five additional topics for inclusion in round two (Supplementary Table 2); none were proposed by two or more participants, so these were not added for round 2 based on our study criteria. Round one data were analysed descriptively (Stata version 17.0).

Participants were then invited to round two, which included the same topics. For each topic in round two, participants were shown the average round one ratings from both participant groups and their own previous score and could retain or revise their rating. The same approach was used in round three, which included only topics not reaching consensus in round two, based on pre-specified definitions as outlined in our protocol:^23^

1. ‘Consensus in’: ≥70% of participants in both groups score the topic ‘Critically important’ and ≤15% of participants in both groups score the topic ‘No or limited importance’
2. ‘Consensus out’: ≥70% of participants in both groups score the topic ‘No or limited importance’ and ≤15% of participants in both groups score the topic ‘Critically important’
3. ‘No consensus’: anything else

All surveys were administered using DelphiManager, a web-based software system designed to create, manage and analyse multi-round Delphi surveys.^26^

### Step-3: Consensus workshop

All participants who indicated in round three that they were interested in taking part in a consensus workshop were invited. Based on availability, an initial one-hour online meeting was held in March 2025 with seven people of reproductive age to support introductions, explain the process, and promote confidence in sharing views. A final three-hour online consensus workshop took place in April 2025.

Before the workshop, participants received the agenda, biographies, and topic list. Two independent, experienced facilitators co-chaired the workshop. Topics reaching ‘consensus in’ based on round three were discussed first with the full group. This was followed by two breakout sessions to consider ‘no consensus’ topics, with random allocation ensuring equal representation of both participant groups. Voting was conducted using Mentimeter after each breakout session. A final whole-group discussion explored views on the number of priority topics and the clinical application of the priority topic list. The workshop was recorded for notetaking.

### Patient and public involvement

A UK-wide Public Advisory Group of people of reproductive age contributed throughout all study stages via online meetings and written feedback between February 2023 and June 2025, strengthening relevance, clarity, inclusivity, and real-world applicability. The group was established in 2021, with members recruited through local and national community, charitable and support groups to ensure diversity in terms of age, gender, and health and pregnancy experiences.^27^ Two members (ND and LK) provided additional detailed input on draft versions of the study protocol,^23^ and this final results paper. More detailed contributions and reflections are reported in Supplementary Table 3 in line with GRIPP2 guidance.^28^

## Results

### Step-1: Clinical guideline and literature review, workshops and interviews

The review identified 33 topics (Supplementary Table 4). Eighteen topics were identified from the NICE Clinical Knowledge Summary on preconception advice and management,^18^ with a further seven identified from international preconception care guidelines.^25^ The literature search yielded 166 umbrella reviews and systematic reviews, from which eight additional topics were identified.

Two workshops were conducted with people of reproductive age (n=6 and n=9), and interviews were completed with 14 primary care professionals, including GPs and nurses. Participants came from all four UK nations and were diverse in age, gender and ethnicity (Supplementary Table 5). Additional topics were suggested by people of reproductive age (e.g. ‘Neurodiversity’, ‘Gender identity and gender transition’, ‘Known to other health services’) and practitioners (e.g. ‘Pregnancy anxiety’, ‘Living circumstances’, ‘Ethnicity’). One topic was removed (‘Travel to Zika virus affected areas’ was no longer considered relevant), and ‘Financial security’ was combined with ‘Living circumstances’, resulting in 37 topics for the Delphi survey. Descriptions of several topics were refined to emphasise patient experiences and needs beyond medical diagnoses (Supplementary Table 6).

### Step-2: Delphi survey

Eighty-five participants completed round one, and 63 completed all three rounds (74%). Participant characteristics (Table 1) remained broadly consistent across survey rounds (Supplementary Table 7).

**Table 1.** Characteristics of Delphi survey and final consensus workshop participants.

|  | **Delphi survey round 3**<br>**N = 63** |  | **Final consensus workshop**<br>**N = 21** |  |
| --- | --- | --- | --- | --- |
|  | **People of reproductive age**<br>**n = 35** | **Primary care practitioners**<br>**n = 28** | **People of reproductive age**<br>**n = 10** | **Primary care practitioners**<br>**n = 11** |
| **Place of residence, n (%)** |  |  |  |  |
| England | 26 (74.3) | 15 (53.6) | 6 (60.0) | 11 (100.0) |
| Wales | 2 (5.7) | 3 (10.7) | 1 (10.0) | - |
| Scotland | 3 (8.6) | 5 (17.9) | 3 (30.0) | - |
| Northern Ireland | 4 (11.4) | 5 (17.9) | - | - |
| **Age group, n (%)** |  |  |  |  |
| 16-20 years | 2 (5.7) | - | - | - |
| 21-30 years | 9 (25.7) | - | 2 (20.0) | 2 (18.2) |
| 31-40 years | 13 (37.1) | 15 (53.6) | 6 (60.0) | 5 (45.5) |
| 41-50 years | 11 (31.4) | 6 (21.4) | 2 (20.0) | 1 (9.1) |
| 51-60 years | NA | 4 (14.3) | NA | 1 (9.1) |
| 61-70 years | NA | 1 (3.6) | NA | 1 (9.1) |
| Over 70 years | NA | 1 (3.6) | NA | 1 (9.1) |
| Prefer not to answer | - | - | - | - |
| **Gender, n (%)** |  |  |  |  |
| Woman | 24 (68.6) | 20 (71.4) | 6 (60.0) | 9 (81.8) |
| Man | 8 (22.9) | 8 (28.6) | 3 (30.0) | 2 (18.2) |
| Non-binary | 1 (2.9) | - | - | - |
| Transgender | 2 (5.7) | - | 1 (10.0) | - |
| **Ethnicity, n (%)** |  |  |  |  |
| White | 2 (5.7) | 2 (7.1) | 1 (10.0) | - |
| White British | 16 (45.7) | 10 (35.7) | 6 (60.0) | 9 (81.8) |
| Asian | 4 (11.4) | 5 (17.9) | 1 (10.0) | 1 (9.1) |
| Asian British | 4 (11.4) | 4 (14.3) | 1 (10.0) | - |
| Black | 3 (8.6) | 2 (7.1) | - | 1 (9.1) |
| Black British | 2 (5.7) | 1 (3.6) | - | - |
| Mixed | 3 (8.6) | 1 (3.6) | 1 (10.0) | - |
| Other: Arab | 1 (2.9) | 4 (14.3) | - | - |
| **Previously been pregnant or currently pregnant, n (%)** | 17 (48.6) | NA | 7 (70.0) | NA |
| **Pregnancy and parenthood intentions, n (%)** |  |  |  |  |
| Currently trying for a baby | 5 (14.3) | NA | - | NA |
| Would like to try for a baby in the next year or so | 3 (8.6) | NA | 1 (10.0) | NA |
| Would like to try for a baby, but not yet in the next year | 5 (14.3) | NA | 2 (20.0) | NA |
| Would like to try, but | 5 (14.3) | NA | 2 (20.0) | NA |

|  | Delphi survey round 3<br>N = 63 |  | Final consensus workshop<br>N = 21 |  |
| --- | --- | --- | --- | --- |
|  | People of reproductive age<br>n = 35 | Primary care practitioners<br>n = 28 | People of reproductive age<br>n = 10 | Primary care practitioners<br>n = 11 |
| struggling to get pregnant |  |  |  |  |
| Not sure | 3 (8.6) | NA | - | NA |
| Already have children, don't want to have more | 9 (25.7) | NA | 4 (40.0) | NA |
| Don't have children, and do not want them | 3 (8.6) | NA | - | NA |
| Other: already have a child and unable to have more | 1 (2.9) | NA | - | NA |
| Other: hoping to adopt | 1 (2.9) | NA | 1 (10.0) | NA |
| **Long-term health condition, n (%)** |  |  |  |  |
| Physical health condition | 6 (17.1) | NA | - | NA |
| Mental health condition | 6 (17.1) | NA | 1 (10.0) | NA |
| Physical and mental health condition | 5 (14.3) | NA | 2 (20.0) | NA |
| No long-term health condition | 18 (51.4) | NA | 6 (60.0) | NA |
| Prefer not to answer | - | NA | 1 (10.0) | NA |
| **General practice contact, n (%)** |  |  |  |  |
| At least once every year | 31 (88.6) | NA | 7 (70.0) | NA |
| Most years, but not every year | 4 (11.4) | NA | 3 (30.0) | NA |
| **Main profession, n (%)** |  |  |  |  |
| GP | NA | 11 (39.3) | NA | 8 (72.7) |
| Nurse | NA | 4 (14.3) | NA | - |
| Pharmacist | NA | 2 (7.1) | NA | - |
| Midwife | NA | 1 (3.6) | NA | - |
| Health visitor | NA | 2 (7.1) | NA | - |
| Link worker / social prescriber | NA | 2 (7.1) | NA | - |
| Other: practice manager | NA | 2 (7.1) | NA | - |
| Other: GP trainee / junior doctor / GP registrar | NA | 3 (10.7) | NA | 2 (18.2) |
| Other: Public Health Officer | NA | 1 (3.6) | NA | 1 (9.1) |
| **Professional experience, n (%)** |  |  |  |  |
| Less than 5 years | NA | 11 (39.3) | NA | 7 (63.6) |
| 5-10 years | NA | 5 (17.9) | NA | 1 (9.1) |
| 10-15 years | NA | 3 (10.7) | NA | 1 (9.1) |
| 15+ years | NA | 9 (32.1) | NA | 2 (18.2) |

NA, not applicable, i.e. question not asked.

Eleven topics reached consensus for inclusion after round two, with no additional topics reaching consensus in round three (Table 2, Supplementary Table 8). Differences between participant groups showed that ≥70% of people of reproductive age, but not primary care professionals, scored ‘Infertility issues and treatment’, ‘Mental health’, ‘Genetic conditions’, ‘Infectious diseases’ and ‘Access to care’ as critically important in at least one round. Conversely, ≥70% of primary care professionals, but not people of reproductive age, scored ‘Weight status’, ‘Nutritional deficiencies’, ‘Sexually transmitted infections’, and ‘Folic acid supplement use’ as critically important. For participants who dropped out, scores from their most recent round were retained; conclusions did not differ when compared with scores from participants who completed all three rounds.

**Table 2.** Summary of scoring of topics in the Delphi survey rounds and final consensus workshop.

| Topic | Delphi survey round 1 | Delphi survey round 2 | Delphi survey round 3 | Final consensus workshop |
| --- | --- | --- | --- | --- |
| **Domain 1: future pregnancy plans and previous experiences** |  |  |  |  |
| Pregnancy intention and timing | No consensus | No consensus | No consensus | Include |
| Contraception use | No consensus | No consensus | No consensus | No consensus |
| Pregnancy anxiety | No consensus | No consensus | No consensus | Exclude |
| Infertility issues and treatment | No consensus<sup>a</sup> | No consensus<sup>a</sup> | No consensus | No consensus |
| Previous pregnancy complications and experiences | Include | Include | NA | Include |
| Previous pregnancy or baby loss | Include | Include | NA | Include |
| **Domain 2: medical conditions and tests** |  |  |  |  |
| Mental health | No consensus<sup>a</sup> | Include | NA | Include |
| Physical health | Include | Include | NA | Include |
| Neurodiversity | No consensus | No consensus | No consensus | Exclude |
| Medication use | Include | Include | NA | Include |
| Gender identity and gender transition | No consensus | No consensus | No consensus | Exclude |
| Genetic conditions | No consensus<sup>a</sup> | Include | NA | Include |
| Weight status | No consensus<sup>b</sup> | No consensus<sup>b</sup> | No consensus | Exclude |
| Previous bariatric surgery | No consensus | No consensus | No consensus | Exclude |
| Nutritional deficiencies | No consensus | No consensus<sup>b</sup> | No consensus | Exclude |
| Dental health | No consensus | No consensus | No consensus | Exclude |
| Cervical smear status | No consensus | No consensus | No consensus | Exclude |
| Immunisation status | No consensus | No consensus | No consensus | Include |
| Infectious diseases | No consensus<sup>a</sup> | No consensus | No consensus<sup>a</sup> | Exclude |
| Sexually transmitted infections | No consensus<sup>b</sup> | Include | NA | Include |
| Other health services | No consensus | No consensus | No consensus | Exclude |
| **Domain 3: health behaviours** |  |  |  |  |
| Dietary habits | No consensus | No consensus | No consensus | Exclude |
| Folic acid supplement use | No consensus<sup>b</sup> | No consensus<sup>b</sup> | No consensus<sup>a</sup> | Include |
| Dietary supplement use | No consensus | No consensus | No consensus | Exclude |
| Physical activity level | No consensus | No consensus | No consensus | Exclude |
| Smoking status | Include | Include | NA | Include |
| Alcohol consumption | Include | Include | NA | Include |
| Drug use | Include | Include | NA | Include |
| **Domain 4: social factors and personal circumstances** |  |  |  |  |
| Living circumstances and financial security | No consensus | No consensus | No consensus | Exclude |
| Physical, sexual and emotional abuse | Include | Include | NA | Include |
| Social support | No consensus | No consensus | No consensus | Exclude |
| Partner support and relationship | No consensus | No consensus | No consensus | Exclude |
| Access to care | No consensus | No consensus<sup>a</sup> | No consensus | Exclude |
| Knowledge of preconception health and pregnancy | No consensus | No consensus | No consensus | Include |
| Ethnicity | No consensus | No consensus | No consensus | Exclude |
| **Domain 5: environmental factors** |  |  |  |  |
| Exposure to air pollution | No consensus | No consensus | No consensus | Exclude |
| Exposure to dangerous substances or radiation | No consensus | No consensus | No consensus | Exclude |

<sup>a</sup> topic was scored 'critically important' by $\geq 70\%$ of people of reproductive age but not $\geq 70\%$ of primary care practitioners
<sup>b</sup> topic was scored 'critically important' by $\geq 70\%$ of primary care practitioners but not $\geq 70\%$ of people of reproductive age

### Step-3: Consensus workshop

Twenty-one participants attended the final consensus workshop (Table 1). During the first breakout session, participants discussed 26 topics that had not reached consensus. Three priority topics (‘Weight status’, ‘Folic acid supplement use’, and ‘Patient knowledge of preconception health and pregnancy’) were subsequently voted ‘consensus in’, while two topics were voted ‘consensus out’ (‘Exposure to air pollution’, ‘Exposure to dangerous substances or radiation’) (Supplementary Table 9).

In the second breakout session, 21 remaining ‘no consensus’ topics were discussed. Final voting led to two further topics being prioritised (‘Pregnancy intention and timing’, and ‘Immunisation status’) (Supplementary Table 10). All other topics were excluded, with two reaching ‘no consensus’ (‘Contraception use’, and ‘Infertility issues and treatment’).

During the concluding whole-group discussion, participants agreed that 16 topics were too many to constitute a practical priority list for initial preconception care discussion, and that a shorter list (e.g. a top five) would be more feasible. Following the workshop, online meetings with two GP participants who were interested in supporting the next step informed the initial consolidation of the 16 priority topics into five broader topic areas: ‘Patient knowledge of preconception health and pregnancy’, ‘Ideas, concerns and expectations’, ‘Health conditions’, ‘Health behaviours’ and ‘Physical, sexual and emotional abuse’. These were shared with all consensus workshop participants for feedback and/or agreement.

Based on email responses from 18 participants, the topic ‘Physical, sexual and emotional abuse’ was combined with ‘Mental health’ (under overall topic area ‘Health conditions’). There was general agreement on all other aspects of the list, and for the four priority topic areas with underlying topics as agreed during the consensus meeting to be the final output from the study (Box 2).

## Discussion

### Summary

This study used a multi-stage consensus process to prioritise topics for discussion during opportunistic preconception care in general practice, integrating the perspectives of people of reproductive age and primary care practitioners in the UK. From an initial list of 37 evidence-based topics, four overarching priority areas were identified to guide preconception care discussions: ‘Patient knowledge of preconception health and pregnancy’, ‘Ideas, concerns and expectations’, ‘Health conditions’, and ‘Health behaviours’. These priority areas provide the basis of pragmatic guidance for opportunistic conversations according to patient needs, with the potential to deliver more consistent and feasible preconception care in routine general practice.

### Comparison with existing literature

The priority topics identified in this study broadly align with existing preconception care guidance in the UK and internationally.^18, 25^ However, rather than providing a long, extensive checklist, topics were prioritised and grouped to offer a more structured and flexible approach that can be adapted to the time available and relevance to individual patients.

Many of the topics covered in the NICE Clinical Knowledge Summary on preconception advice and management are represented within our four priority areas.^18^ However, some NICE-recommended items were not prioritised in the final list. These were generally topics considered less suitable for initial, time-limited discussions (e.g. ‘dietary habits’) or areas already covered through routine care (e.g. ‘cervical smear status’).

Some patient-facing resources already exist, such as the FIGO (International Federation of Gynaecology and Obstetrics) preconception checklist which offers comprehensive screening structured around nutrition, supplementation, lifestyle, vaccination and medical conditions.^29^ While evidence-based, its length and level of detail may limit routine use in opportunistic consultations.^30^ Tommy’s pregnancy planning tool was frequently mentioned by GPs in this study as familiar and practical, particularly as it can be completed outside a consultation.^31^ However, such tools rely on patient awareness or health professional signposting, and do not address sensitive topics such as ‘physical, sexual and emotional abuse’ or medical history details such as ‘medication use’ that were prioritised in this study.

A priority list of “red flags” for preconception care has previously been proposed in a clinical review.^8^ While most topics overlap with our findings, the “red flags” do not explicitly include ‘Patient knowledge of preconception health and pregnancy’ and ‘Ideas, concerns and expectations’. Findings from our study suggest these areas are critical, acting as umbrella topics that can quickly identify information gaps, misconceptions, and the need for further support.

We observed differences in topic prioritisation between people of reproductive age and primary care practitioners. These may reflect variations in personal experience, for example, related to the importance of emotional support when struggling to conceive. Differences may also reflect levels of knowledge about preconception health. For example, agreement on the importance of folic acid supplement use and immunisation was only achieved based on discussion during the final consensus workshop. These findings are consistent with existing literature highlighting the need to address gaps in knowledge and awareness on preconception health among people of reproductive age.^4, 32–34^

The topic of ‘ethnicity’ was discussed extensively in the consensus workshop but not prioritised in the final list, despite strong evidence linking ethnic background to maternity outcomes and health disparities.^35^ This may reflect uncertainty about how best to address ethnicity within brief consultations, rather than a lack of perceived importance. Consensus workshop participants highlighted its relevance for tailoring advice, identifying genetic risks, and ensuring early access to antenatal care and culturally appropriate communication. In line with NHS recommendations,^36^ these findings suggest further research is needed on how sociodemographic and social factors such as ethnicity can be incorporated sensitively and meaningfully within conversations in primary care.

### Strengths and limitations

Strengths include the integration of perspectives of people of reproductive age and primary care practitioners using a rigorous, multi-stage consensus methodology, underpinned by strong patient and public involvement. Potential topics were derived from current NICE guidance, international recommendations and recent literature. The explicit focus on feasibility in prioritising topics enhances applicability to routine general practice.

The study also included a diverse UK-wide participant sample in terms of age and ethnicity, pregnancy intentions and health conditions (people of reproductive age) and professional experience (practitioners). However, most Delphi survey participants were women, and the practitioners in the final consensus workshop were all GPs from England, which may have influenced prioritisation. Additional limitations include potential participant self-selection bias, the UK-specific context, and the use of a hypothetical scenario that may not fully capture the complexity of real-world consultations.

### Implications for policy, practice and research

This study identified several preconception care-relevant topics not currently included in NICE guidance,^18^ most notably ‘patient knowledge of preconception health and pregnancy’, ‘previous pregnancy or baby loss’, ‘sexually transmitted infections’, and ‘physical, sexual and emotional abuse’. These topics were identified in early qualitative stages of this study, and prioritised through the consensus process, suggesting they warrant consideration in future guidance.

For clinical practice, resources and systems are needed to support routine use of the priority list, for example during contraception and medication reviews, or when promoting healthy weight or smoking cessation.^19, 37–39^ Embedding the priority list within a preconception care pathway could facilitate opportunistic delivery, for example using a modified Ask-Advice-Assist approach: brief screening aligned with the prioritised areas (Ask), initial advice and signposting (including digital tools) (Advice), and follow-up where appropriate (Assist). Integration within electronic medical records, feeding into a preconception care template, alongside health professional education, may further enhance feasibility and identification of patients’ preconception care needs.^6, 7, 11, 40–42^ Further research should co-develop comprehensive preconception care pathways that are acceptable to patients and meet practitioners’ training and resource needs, and evaluate impacts on health, pregnancy outcomes and disparities.

### Conclusion

This study provides a consensus-based, prioritised framework for preconception care discussions in general practice that balances evidence, feasibility and patient-centredness. Identifying four overarching prioritised areas, it offers a practical alternative to lengthy checklists and supports a flexible approach to opportunistic preconception care. Integration into guidance and primary care systems has the potential to improve consistency and quality of preconception care, with important implications for maternal, perinatal and long-term health outcomes.

## Data Availability

All data produced in the present study are available upon reasonable request to the authors.

## Additional information

### Ethical approval

This study has been approved by the University of Southampton Faculty of Medicine Ethics Committee (ERGO 83699 and 92950).

## Acknowledgements

The authors would like to thank members of the Project Advisory Group and the Public Advisory Group for their valuable contributions to the development of the study protocol and for their input into discussions on the implications of the findings. We extend a particular thank you to members of the Public Advisory Group who provided detailed feedback on study materials and pilot tested the survey: Nathan, Laura, Khadija, Kate, Sarah, Megan, Michelle, Tanjida and Rosie. We are also grateful to organisations and charities supporting participant recruitment by sharing information about the study, including the Primary Care Women’s Health Society (PCWHS), GPs Championing Perinatal Care (GPCPC), WiseGP, Society for Academic Primary Care (SAPC), Sands, Beyond Reflections and Active Pregnancy Foundation. We thank all participants who took part at each stage of the study for their time, insights, and willingness to share personal reflections and experiences. Finally, we would like to thank Nahid Ahmad and Louise Dunford for co-facilitating the final consensus workshop, and Isla Davis for supporting participants throughout the workshop.

## Boxes

**Box 1.**
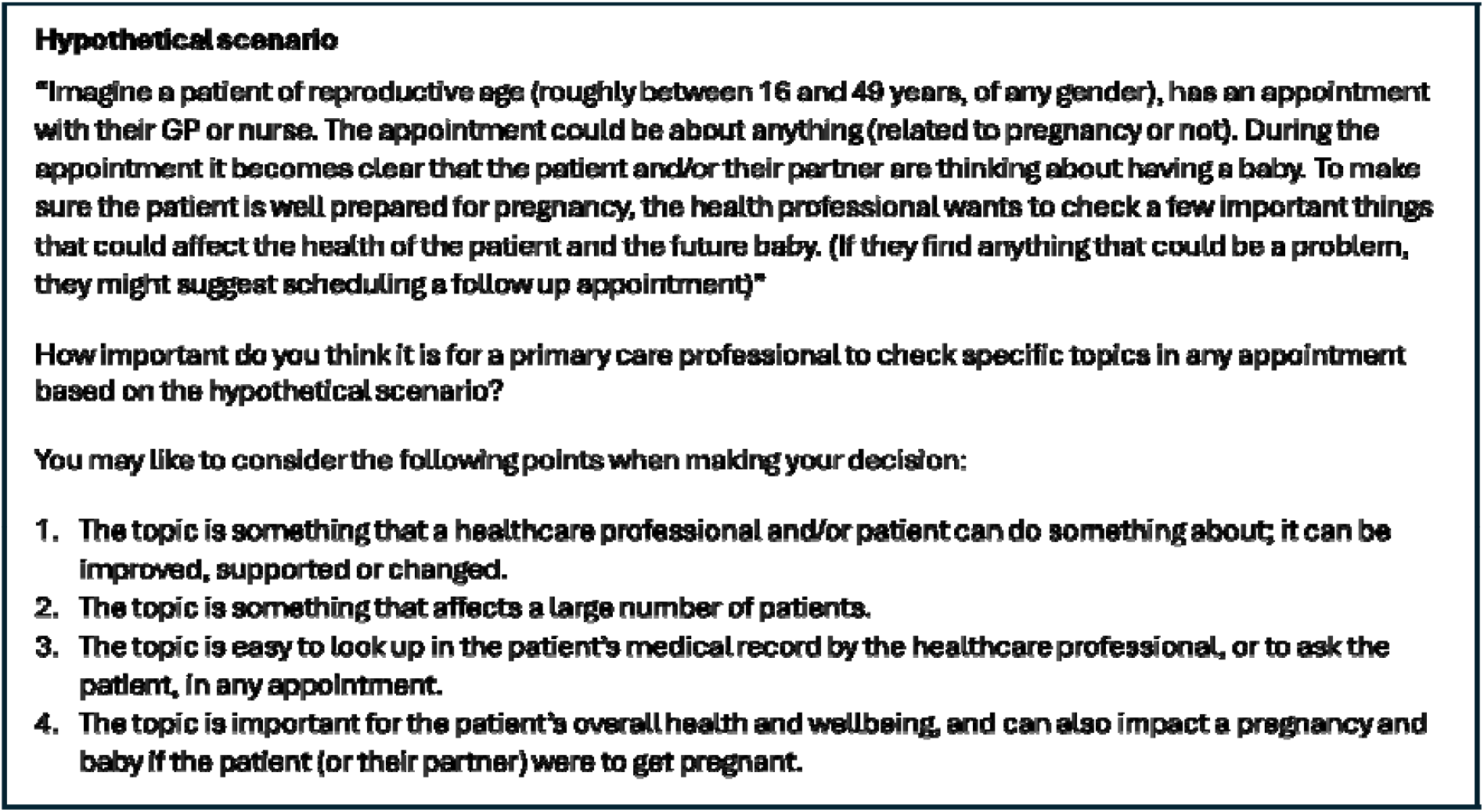
Hypothetical scenario and prioritisation principles presented to Delphi survey participants.

**Box 2.**
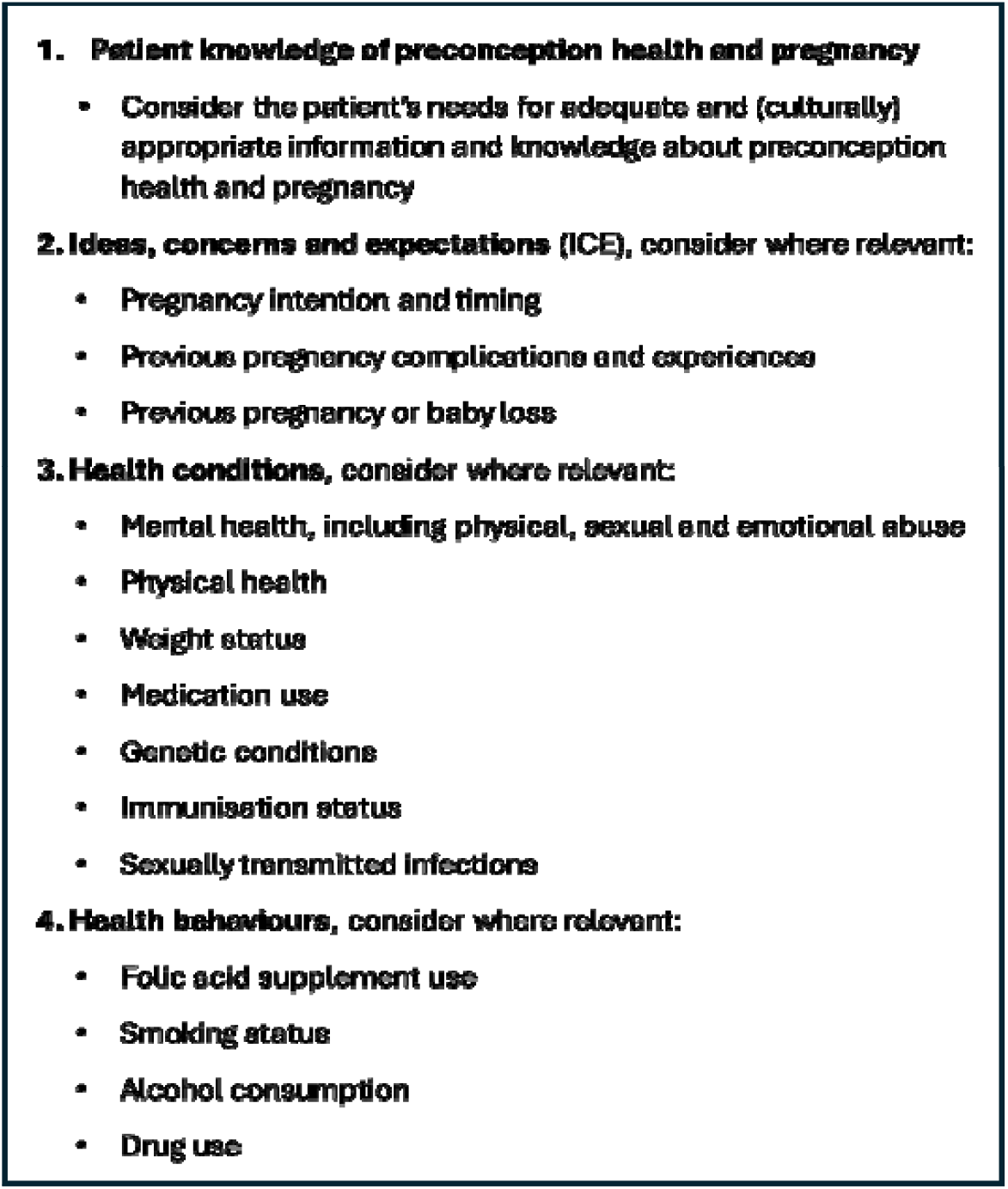
Agreed priority topic areas to guide a structured and flexible approach to opportunistic preconception care in general practice.

