## Supplement for "Priority topics for preconception care in general practice: a consensus study"

### Supplementary Tables

**Supplementary Table 1.** Literature search to identify umbrella reviews and systematic reviews of studies on preconception health published in English between 1 November 2018 - 30 November 2023

| **Database** | **Search terms** | **Number of records identified** |
| --- | --- | --- |
| CINAHL | TI preconception OR TI pre-conception OR AB preconception OR AB pre-conception OR TI prepregnancy OR TI pre-pregnancy OR AB prepregnancy OR AB pre-pregnancy  Restricted to: English language; Peer-Reviewed; Human; Systematic Review | 92 |
| OVID MEDLINE | (preconception or pre-conception or prepregnancy or pre-pregnancy).ti. and (preconception or pre-conception or prepregnancy or pre-pregnancy).ab.  Restricted to: English language; Review articles | 93 |
| EMBASE | (preconception.ti. or preconception.ab. or pre-conception.ti. or pre-conception.ab. or prepregnancy.ti. or prepregnancy.ab. or pre-pregnancy.ti. or pre-pregnancy.ab.) and (review.ti. or review.ab.)  Restricted to: English language; Human | 1185 |
| Total |  | 1370 |
| Duplicates |  | 146 |
| Total after duplicates removed |  | 1,224 |
| Not relevant |  |  |
| Studies included |  | 166 |

**Supplementary Table 2.** Additional topic suggestions in round one

| **Additional topic suggestions** | **Participant group** |
| --- | --- |
| “Female genital mutilation” | Person of reproductive age |
| “Fertility chances - Timing; chances of getting pregnant now vs in the future; options around fertility preservation or going ahead” | Person of reproductive age |
| “Are they a vulnerable adult; for example; have a learning disability? (to enable support to be put in place; if needed)” | Primary care professional |
| “Do they have certain religious beliefs that might impact on health beliefs and practices; like Jehovah's Witnesses and no blood transfusions? (so that it can be noted for future reference and a discussion can be had before it gets into a hot emotive situation)” | Primary care professional |
| “Do they have any strongly held health beliefs that might diverge from standard practice; like anti-vaccine; natural birthing? (for reference and current/future discussion)” | Primary care professional |

**Supplementary Table 3**. Patient and public involvement throughout the study, reported in line with the GRIPP2 short form^a^

| **Aim** | Patient and public involvement (PPI) aimed to ensure that the research was relevant, inclusive, and informed by the perspectives and lived experiences of people of reproductive age in the UK. PPI sought to refine the study aims, improve the clarity and accessibility of study materials, support inclusive recruitment and engagement of study participants, and ensure that consensus-building processes and dissemination activities reflected patient and public priorities and preferences. |
| --- | --- |
| **Methods** | A Public Advisory Group including people of reproductive age from across the UK were involved through online group meetings. The group was established in 2021, with members recruited through local and national community, charitable and support groups to ensure diversity in terms of age, gender, and health and pregnancy experiences. Seven 1-hour meetings took place between February 2023 and June 2025, attended by on average 15 public contributors. Two members of the Public Advisory Group - Nathan Davies and Laura Kubelabo - provided further, more detailed input through written feedback on draft versions of the study protocol and final results paper – which were also discussed in online meetings in August 2023 and March 2026. Nathan and Laura, alongside public contributors Khadija, Kate, Sarah, Megan, Michelle, Tanjida and Rosie, also provided detailed written feedback and suggestions on study participant documents. All public contributors were reimbursed for their time through Amazon e-vouchers, in line with NIHR guidance. |
| **Results** | Outcomes of including PPI in the study include:   - *Protocol development*: Patient and public involvement led to simplification and clarification of the study aim and improved explanation of key concepts and context in the protocol. PPI confirmed the relevance of the study to people of all gender identities and informed the use of inclusive language relating to ‘preparing for pregnancy and parenthood’. Early involvement also clarified the study scope and established meaningful opportunities for public involvement and participation across all study stages. - *Participant information*: Public contributors (Nathan, Laura, Khadija, Kate, Sarah, Megan, Michelle, Tanjida, and Rosie) reviewed and suggested improvements to all participant-facing study documents. Their feedback led to recruitment materials being shortened and written in clearer, more accessible language, making the study easier to understand and more inviting to potential participants. They also advised clarifying the timing and purpose of different survey rounds, which improved transparency and helped manage participant expectations. In addition, public contributors highlighted the importance of clearly stating reimbursement arrangements; as a result, this information was made more prominent and consistently presented across all documents. - *Topic guides and workshop content*: Public contributors informed the development of workshop topic guides and facilitation approaches to ensure inclusive, supportive, and meaningful participation (Step 1 and 3). They advised on setting clear expectations and ground rules, using introductions and icebreakers, and creating multiple opportunities for contribution, including free discussion, turn-taking, and the option to pass. PPI input shaped the use of breakout groups, pre-meeting briefings, and facilitation strategies to build confidence and support participation, particularly for those who may feel less confident speaking in larger groups. - *Study participant recruitment*: Public contributors shaped a flexible and inclusive recruitment strategy by recommending recruitment through multiple settings and platforms. Several meetings involved discussions around recruitment of diverse study participants, including groups of people to approach, how to do this, and how to improve recruitment approaches for Step 2 based on experience of reaching out to personal networks for Step 1 recruitment.   Groups of people to approach: Public contributors suggested reaching out to their friends, colleagues and community connections (e.g. through sports clubs and schools). They also supported targeted outreach to underserved groups, including younger people, people from ethnic minority backgrounds, non-binary and transgender individuals, and people living across all four UK nations.  How to approach: Public contributors suggested using multiple ways to reach out to potential study participants, including social media, WhatsApp messages, word of mouth, and used study-provided templates alongside personalised messages.  Learnings: Based on reflections after Step 1, public contributors identified improvements to enhance study participant recruitment for Step 2, including broadening outreach beyond health-interested connections and networks, personalising invitations and sharing own experiences with being involved in research, following up with contacts, and supporting participation for people with limited internet access or English proficiency.   - *Dissemination*: Public contributors shaped a dissemination plan to ensure study findings reach the public, including underserved communities. They discussed that sharing findings would have multiple goals: to increase knowledge, raise awareness, and normalise discussion about the importance of being as healthy as possible before pregnancy for people of all genders; factors affecting conception, pregnancy, and baby health; opportunities to seek advice and support from health professionals; and the key topics that might be discussed. Contributors suggested accessible formats (blogs, infographics, lay summaries, videos) and multiple platforms (social media, GP practices, schools, pharmacies, community and third-sector organisations), emphasising translation, easy-to-read language, and targeted outreach. |
| **Discussion** | Public contributors shaped all study stages, enhancing the study’s overall relevance, clarity, inclusivity, and real-world applicability. They refined the aim, reviewed materials, piloted the survey, guided workshops, informed recruitment to reach diverse groups, and co-developed accessible dissemination plans. Collectively, this will help raise awareness and normalise discussion about pregnancy preparation in primary healthcare encounters.  Public contributors shared their take-away messages from the study findings, things they learned from the study that they would tell their friends:   - “It’s helpful to talk to a doctor or nurse early if you’re thinking about having a baby, even if it’s not right away. Small steps like taking folic acid, stopping smoking and checking your physical and mental health can really help.” - “You need to think about health before pregnancy long before you are even ready for trying. Try to become the healthiest version of yourself before trying.” - “Preconception health is important for parent and child – not just when it’s a baby but also as they grow older.” - “It is important to recognise that many factors impact and influence preconception health.” - “There is something coming about preconception health. You have to ask your GP. If you have any concern, don’t feel shy.” - “Preconception health is important, and communication influences how diverse audiences are reached.” - “I’d tell everyone of suitable age to be aware that there could be a comprehensive infrastructure (soon to be built) in which a GP can advise you if you’re considering having a baby!” |
| **Reflections and critical perspectives** | Public contributor co-authors Nathan and Laura shared their reflections.  Things that went will include:   - *Early and meaningful involvement*: PPI was embedded from the outset, ensuring the study was relevant to people of reproductive age. Contributions have been genuinely considered and incorporated, demonstrating meaningful rather than tokenistic involvement. - *Inclusive and well-facilitated engagement*: Meetings have been well organised and facilitated, with clear agendas, reminders, and opportunities for all members to contribute. - *Positive and supportive group environment***:** The PPI group is well established, with a friendly and supportive atmosphere that encourages open participation. Continued engagement from members suggests the group feels valued and involved. - *Efficient reimbursement processes***:** Payments for PPI activities have been prompt, straightforward, and aligned with NIHR rates.   Things that could be improved include:   - *Opportunities for in-person engagement***:** While online meetings have been convenient for many, offering occasional in-person or hybrid options may benefit some members. - *Training and development for PPI members***:** Some participants may have welcomed optional training or development opportunities to support their role in the project. - *Broader age representation***:** Although the group was diverse, including more younger members could better reflect the full age range of people of reproductive age. - *Public communication and visibility***:** A project website or social media presence could help share ongoing updates, progress, and next steps with the wider public. - *Ongoing feedback processes***:** Establishing a simple log for PPI members to record reflections throughout the project could make it easier to capture what is working well and what could improve, supporting continuous rather than end-of-project feedback. |

^a^ Staniszewska S, Brett J, Simera I, Seers K, Mockford C, Goodlad S, et al. GRIPP2 reporting checklists: tools to improve reporting of patient and public involvement in research. BMJ. 2017;358:j3453.

**Supplementary Table 4.** Preconception health topics identified through clinical guidelines (UK NICE Clinical Knowledge Summary [NICE CKS] and a systematic review of international preconception care guidelines [Dorney et al]) and additional literature reviews

| **Topics** | **Example screening question based on clinical recommendation/intervention** | **Source** | **Population** |
| --- | --- | --- | --- |
| **Section 1: Future pregnancy plans and previous and experiences** | |  |  |
| Pregnancy intention and timing | When is the patient planning to start trying to conceive (again)? | NICE CKS | Female & male |
| Contraception use | Is the patient ready to start trying to conceive now, or would they need (advice on) effective contraception to reduce the risk of an unplanned pregnancy? | NICE CKS | Female & male |
| Infertility issues and treatment | Does the patient experience infertility, and have they used or are they currently undergoing infertility treatment? | Jenabi et al. | Female & male |
| Previous pregnancy complications | Has the *female* patient experienced complications, such as gestational diabetes or pre-term birth, in a previous pregnancy? | NICE CKS | Female |
| Previous pregnancy loss | Has the patient previously experienced pregnancy loss? | Patel et al. | Female & male |
| **Section 2: Health behaviours** | |  |  |
| Dietary habits | Is the patient consuming a healthy and balanced diet? | NICE CKS | Female & male |
| Folic acid supplementation | Is the *female* patient taking the appropriate daily dose of folic acid? | NICE CKS | Female |
| Dietary supplement use | Is the patient using any dietary supplements (other than folic acid)? | Dorney et al. | Female & male |
| Physical activity level | Does the patient engage in regular, moderate-intensity physical activity or exercise? | Dorney et al. | Female & male |
| Smoking status | Does the patient smoke? | NICE CKS | Female & male |
| Alcohol consumption | Does the patient consume alcohol? | NICE CKS | Female & male |
| Recreational or illicit drug use | Does the patient use recreational or illicit drugs? | NICE CKS | Female & male |
| **Section 3: Medical conditions** | |  |  |
| Mental health conditions | Does the patient have a history or current mental health condition, and how is this currently treated and managed? | NICE CKS | Female & male |
| Physical health conditions | Does the patient have a chronic medical condition or disability, and how is this currently treated and managed? | NICE CKS | Female & male |
| Dental health | Does the patient have good oral health and have they visited a dentist in the past year? | Zaçe et al. | Female & male |
| Medication use | Is the patient taking prescription medications, over-the counter medicines or herbal remedies that may influence fertility or are not recommended for use in pregnancy? | NICE CKS | Female & male |
| Genetic conditions | Is the patient at risk of having a baby with an inherited genetic disorder based on their personal, partner or family history? | NICE CKS | Female & male |
| Weight status | Is the patient’s weight in the healthy range based on their body mass index? | NICE CKS | Female & male |
| Bariatric surgery | Has the patient had bariatric surgery? | Caut et al. | Female & male |
| Nutritional deficiencies | Does the patient have, or are they at risk of, nutritional deficiencies (such as iron, iodine and vitamin D)? | Dorney et al. | Female & male |
| **Section 4: Screening, immunization and infections** | |  |  |
| Cervical smear status | Is the *female* patient up-to-date with their cervical smear test (if relevant)? | NICE CKS | Female |
| Immunization status | Is the *female* patient up-to-date with their immunization (for example, are they protected against measles, mumps and rubella [MMR], varicella [chickenpox] and diphtheria-tetanus-pertussis [DTP])? | NICE CKS | Female |
| Infectious diseases | Does the patient have, or are they at risk of, an infectious disease (such as hepatitis B, hepatitis C or HIV)? | NICE CKS | Female & male |
| Sexually transmitted infections (STIs) | Does the patient have a sexually transmitted infection? | Dorney et al. | Female & male |
| Plans to travel to an area with active Zika virus transmission | Is the patient planning to travel to an area with active Zika virus transmission? | NICE CKS | Female & male |
| **Section 5: Psychosocial factors** | |  |  |
| Financial security | Does the patient have access to adequate financial resources? | Dorney et al. | Female & male |
| Physical, sexual and emotional abuse | Does the patient have a past or current experience, or are they at risk, of physical, sexual or emotional abuse (including adverse childhood experiences, intimate partner violence and coercion)? | Dorney et al. | Female & male |
| **Section 6: Access to care, information and support** | |  |  |
| Access to care | Does the patient have access to relevant and appropriate health and community care to be well supported before, during and after pregnancy? | Dorney et al. | Female & male |
| Knowledge of preconception health and pregnancy | Does the patient have adequate and (culturally) appropriate (access to) information and knowledge about preconception health and pregnancy? | Welshman et al. | Female & male |
| Social support | Does the patient have adequate social support before, during and after pregnancy, for example through a network of family and friends? | Kandel et al. | Female & male |
| Partner support | Does the patient feel they have adequate support from their partner before, during and after pregnancy? | Welshman et al. | Female & male |
| **Section 7: External factors** | |  |  |
| Air pollution | Is the patient exposed to high levels of air or traffic pollution? | Caut et al. | Female & male |
| Exposure to hazardous substances and radiation | Is the patient exposed to toxic substances or chemicals in their home, workplace or surrounding environment? | NICE CKS | Female & male |

Caut C, Schoenaker D, McIntyre E, Vilcins D, Gavine A, Steel A. Relationships between Women's and Men's Modifiable Preconception Risks and Health Behaviors and Maternal and Offspring Health Outcomes: An Umbrella Review. Semin Reprod Med. 2022;40(3-04):170-183.

Dorney E, Boyle JA, Walker R, Hammarberg K, Musgrave L, Schoenaker D, Jack B, Black KI. A Systematic Review of Clinical Guidelines for Preconception Care. Semin Reprod Med. 2022;40(3-04):157-169.

Jenabi E, Salimi Z, Ayubi E, Bashirian S, Salehi AM. The environmental risk factors prior to conception associated with placental abruption: an umbrella review. Syst Rev. 2022;11(1):55.

andel P, Lim S, Pirotta S, Skouteris H, Moran LJ, Hill B. Enablers and barriers to women's lifestyle behavior change during the preconception period: A systematic review. Obes Rev. 2021;22(7):e13235.

Patel K, Pirie D, Heazell AEP, Morgan B, Woolner A. Subsequent pregnancy outcomes after second trimester miscarriage or termination for medical/fetal reason: A systematic review and meta-analysis of observational studies. Acta Obstet Gynecol Scand. 2024;103(3):413-422.

Welshman H, Dombrowski S, Grant A, Swanson V, Goudreau A, Currie S. Preconception knowledge, beliefs and behaviours among people of reproductive age: A systematic review of qualitative studies. Prev Med. 2023;175:107707.

Zaçe D, Orfino A, Mariaviteritti A, Versace V, Ricciardi W, DI Pietro ML. A comprehensive assessment of preconception health needs and interventions regarding women of childbearing age: a systematic review. J Prev Med Hyg. 2022;63(1):E174-E199.

**Supplementary table 5.** Characteristics of participants who took part in Step 1 workshops (people of reproductive age) and interviews (primary care practitioners)

|  | **People of reproductive age N = 15** | **Primary care practitioners N = 14** |
| --- | --- | --- |
| **Place of residence, n** |  |  |
| England | 10 | 6 |
| Wales | 1 | 3 |
| Scotland | 3 | 1 |
| Northern Ireland | 1 | 4 |
| **Age group, n** |  |  |
| 21-30 years | 7 | 2 |
| 31-40 years | 5 | 6 |
| 41-50 years | 3 | 2 |
| 51-60 years | *NA* | 2 |
| 61-70 years | *NA* | 2 |
| **Gender, n** |  |  |
| Woman | 8 | 11 |
| Man | 5 | 3 |
| Non-binary | 1 | - |
| Other: Transmasculine | 1 | - |
| **Ethnicity, n** |  |  |
| White | 3 | 2 |
| White British | 4 | 6 |
| Asian | 2 | - |
| Asian British | - | 3 |
| Black British | 1 | - |
| Mixed | 3 | 2 |
| Other: Asian Irish | - | 1 |
| Other: Latino British | 1 | - |
| Other: Hispanic | 1 | - |
| **Long-term health condition, n** |  |  |
| Physical health condition | 3 | *NA* |
| Mental health condition | 3 | *NA* |
| No long-term health condition | 9 | *NA* |
| **General practice contact, n** |  |  |
| At least once every year | 10 | *NA* |
| Most years, but not every year | 4 | *NA* |
| Not every year | 1 | *NA* |
| **Main profession, n** |  |  |
| GP | *NA* | 6 |
| Practice Nurse | *NA* | 2 |
| Other: Advanced Nurse Practitioner | *NA* | 5 |
| Other: Public Health Officer | *NA* | 1 |
| **Professional experience, n** |  |  |
| Less than 5 years | *NA* | 5 |
| 5-10 years | *NA* | 4 |
| 10-15 years | *NA* | 1 |
| 15+ years | *NA* | 4 |
| **Special interest, n** (multiple answer options possible) |  |  |
| Women’s health | *NA* | 10 |
| Sexual health | *NA* | 10 |
| Reproductive health | *NA* | 6 |
| Obstetrics and gynaecology | *NA* | 4 |
| Preconception health | *NA* | 4 |
| Other: Diabetes | *NA* | 2 |
| **Academic role, n** |  |  |
| No | *NA* | 12 |
| Yes, also employed as academic | *NA* | 2 |

*NA*, not applicable, i.e. question not asked.

**Supplementary table 6.** Preconception health topics and example questions identified from literature and clinical guideline review, with changes made based on workshops with people of reproductive age and interviews with primary care practitioners in Step 1 of the study.

| **Preconception health topics** | **Example screening questions for primary care practitioners to consider** | **Supporting quote(s) based on workshops and interviews** |
| --- | --- | --- |
| **Domain 1: Future pregnancy plans and previous experiences** | | |
| Pregnancy intention and timing | When is the patient planning to start trying to conceive (again)? *Is there enough time to optimise risk factors, and should age be discussed?* | “I think the context is also important, like, potentially they're not thinking about children at the moment, but maybe that's something they want in the future, and they want to know when to plan to make sure the timing is right. Kind of like what age should they be thinking about. I'm just thinking of quite a few media stories at the moment, around people in their 40s kind of being worried about fertility.” (PRA) |
| Contraception use | Is the patient ready to start trying to conceive now, or would they need (advice on) effective contraception to reduce the risk of an unplanned pregnancy? | - |
| *Pregnancy anxiety* (newly added) | *What is the plan on how to get pregnant? Is the patient anxious or nervous about becoming pregnant, how do they plan to get pregnant and how do they think they will cope?* | “If someone indicates they want to get pregnant, you know, I then first say what is your plan. Is there anything I can help with or that you’re worried about? Is there anything you know that you are anxious or nervous about? They have often heard lots of stories, so we need to encourage them to explore this, like through ICE model of care which is ideas, concerns and expectations.” (PCP) |
| Infertility issues and treatment | *Does the patient have concerns about their fertility* or are they having difficulty conceiving*? Have they used or are they currently undergoing infertility treatment?* | “When I was younger it would have opened a door if I was asked about any concerns about fertility or something along those lines. I was a bit concerned that I had really heavy periods and I didn’t know whether that's going to affect anything and it might just give the opportunity to screen for those early fertility problems or concerns. If I was asked as a teenager, I would have said actually, yeah, I am worried.” (PRA) |
| Previous pregnancy complications *and experiences* | *What are the patient’s experiences with a previous pregnancy*, including complications such as gestational diabetes, pre-term birth *or caesarean section* (female patients), *antenatal or postnatal mental health issues, or a difficult birth*? | “The psychological impact of pregnancy is really important to consider, you know, previous experiences like pregnancy complications, a difficult or traumatic birth, or having a caesarean section, these have lasting effects on mental wellbeing. Also time spent in hospital and how supported they felt during that experience can also influence mental health, you know, during, but also after that pregnancy. Exploring these experiences helps us understand how past pregnancies may still be affecting them.” (PCP) |
| Previous pregnancy *or baby* loss | Has the patient previously experienced pregnancy *or baby* loss? | “To make this relevant to everyone, you know also thinking about men, I think you need to also talk about baby loss specifically.” (PRA) |
| **Domain 2: Medical conditions and tests** | | |
| Mental health *~~conditions~~* | *How is the patient’s mental health?* Do they have a history of or current mental health condition, and how is this treated and managed *(e.g. potentially teratogenic / harmful medications)? Is there a family* *history of (postpartum) mental health conditions? Does the patient feel they need support with their mental health*? | “Because I think people cannot be in a great place and feel like they need support with their mental health without necessarily already having a label or wanting a label of, you know, depression or anxiety or PTSD or schizophrenia or whatever.” (PRA)  “Could you not just say how is your mental health? You know, do you need any support with your mental health rather than it more rather than it being about a diagnosis?” (PRA) |
| Physical health *~~conditions~~* | *How is the patient’s physical health?* Does the patient have a chronic medical condition or disability, and how is this treated and managed? *(e.g. potentially teratogenic / harmful medications)* | “Same as with mental health.” (PRA) [see topic above] |
| *Neurodiversity* (newly added) | *Is the patient neurodiverse, for example do they experience autism, ADHD or a neurodevelopmental condition?* | “Not sure if it will make the final shortlist, but it may be important if a person is neurodivergent. I had a friend that was diagnosed with autism when she was pregnant. Because hormones just made her really unable to cope, and it really affected her mental health.” (PRA) |
| Medication use | Is the patient taking prescription medication or over-the counter medicines ~~or herbal remedies~~ that may influence fertility or that are not recommended for use before or during pregnancy? | - |
| *Gender identity and gender transition* (newly added) | *Has the patient undergone gender transition? Is the patient receiving any hormone treatment? Have they undergone any surgeries? Are they considering egg freezing?* | “I mean obviously would only apply to a very small minority of patients, but you haven't got anything in here about gender transition. Like is the patient receiving any hormone treatment for gender transition? Have they undergone any surgeries?” (PRA) |
| Genetic conditions | Is the patient at risk of having a baby with an inherited genetic disorder based on their personal, partner or family history? | - |
| Weight status | Is the patient’s weight in the healthy range based on their body mass index? *Do they experience weight issues?* | “And also I think from like friends who would call themselves fat that like when they've seen a GP so much of the pressure is on their weight loss before any other questions about their health, which can be really frustrating for them, yeah, like delegitimising for them, if that makes sense. I understand weight is important, but you want to be as sensitive as possible and not kind of medicalise someone’s weight necessarily. Maybe ask people about weight and also if they think they have weight issues, to make it more personal to talk about?” (PRA) |
| Previous bariatric surgery *(weight loss surgery)* | Has the patient had bariatric surgery *(weight loss surgery)*? | “The only one that I had to Google was about, uh, you know, terms like bariatric surgery.” (PRA) |
| Nutritional deficiencies | Does the patient have, or are they at risk of, nutritional deficiencies (such as iron, iodine or vitamin D)? | - |
| Dental health | Does the patient have good oral health and have they visited a dentist in the past year? | - |
| Cervical smear status | Is the female patient up-to-date with their cervical smear test (if relevant)? | - |
| Immunization status | Is the female patient up-to-date with their immunization. For example, are they protected against measles, mumps and rubella (MMR), varicella (chickenpox) and diphtheria-tetanus-pertussis (DTP)? | - |
| Infectious diseases | Does the patient have, or are they at risk of, an infectious disease (such as hepatitis B, hepatitis C or HIV)? | - |
| Sexually transmitted infections (STIs) | Does the patient have a sexually transmitted infection? | - |
| *Other health services* (newly added) | *Is the patient known to other services (such as social services or mental health services)* | “Is there a question around asking whether the patient is known to other services like, you know mental health services or social services, whatever it is.” (PRA) |
| **Domain 3: Health behaviours** | | |
| Dietary habits | Is the patient consuming a healthy and balanced diet? | - |
| Folic acid supplement use | Is the female patient taking the appropriate daily dose of folic acid? | - |
| Dietary supplement use | Is the patient using any dietary supplements (other than folic acid)? | - |
| Physical activity level | Does the patient engage in regular, moderate-intensity physical activity or exercise? | - |
| Smoking status | Does the patient smoke? | - |
| Alcohol consumption | Does the patient consume alcohol? | - |
| *~~Recreational or illicit~~* Drug use | Does the patient use *~~recreational or illicit~~* drugs *(such as marijuana, cocaine, opioids)?* | “I think the one about drugs needs a bit more, simple detail. I don’t think people can relate to recreational or illicit so maybe leave that out but give some examples, you know like marijuana, if that is what you mean.” (PRA) |
| **Domain 4: Psychosocial factors and personal circumstances** | | |
| Financial security *and living circumstances* | Does the patient *feel they* have access to adequate financial resources? *What are the patient’s living circumstances, such as secure housing, food poverty, fuel poverty, hygiene?* | “I thought the question about adequate financial resources, I think that's a really good question to ask as not all the people think about it necessarily. It’s great this can come from a GP. But I would question what is adequate financial resources and I think that threshold will be very different for everyone. Like what is desired will be quite different in everyone's mind.” (PRA)  “When we were just talking about finances and stuff, maybe housing is important too, maybe whether it's like secure or not.” (PRA)  “Beyond finances, people’s living circumstances, like housing, food poverty, fuel poverty, hygiene you know, that’s gonna have a massive impact on getting pregnant, staying pregnant, all of those things, looking after a child.” (PCP) |
| Physical, sexual and emotional abuse | Does the patient have a past or current experience, or are they at risk, of physical, sexual or emotional abuse (including adverse childhood experiences, intimate partner violence and coercion)? | - |
| Social support | Does the patient have adequate social support before, during and after pregnancy, for example through a network of family and friends, *or do they think they need extra support*? | “I was just thinking about my perspective, what I can do to help my partner or what kind of information I should know to make things better, like it is important to ask maybe if I or my partner need extra support or information.” (PRA) |
| Partner support *and relationship* | *Does the patient have a partner, and do they* feel they have adequate support from their partner before, during and after pregnancy? *Does the patient feel they are in a safe relationship?* | “And I suppose the only other thing I noticed as we were just talking there, is there, you know, are you in a safe relationship or do you feel supported by your partner?” (PRA) |
| Access to care | Does the patient have access to relevant and appropriate health and community care to be well supported before, during and after pregnancy? | - |
| Knowledge of preconception health and pregnancy | Does the patient have adequate and (culturally) appropriate (access to) information and knowledge about preconception health and pregnancy, *or would they like further information*? | “I was just thinking about my perspective, what I can do to help my partner or what kind of information I should know to make things better, like it is important to ask maybe if I or my partner need extra support or information.” (PRA) |
| *Ethnicity* (newly added) | *What influence could the patient’s ethnic background have on their health and wellbeing? Are there any potential barriers to accessing timely and appropriate (maternity) care?* | “Is ethnicity in the list or not? The reason I'd be thinking about this is just because we all know maternal outcomes are worse for some women and I’d want to keep that in the front of my mind to think about what are the potential barriers here for this woman or making sure they are referred to maternity services in a timely manner you know, I think that’s so important to consider.” (PCP) |
| **Domain 5: Environmental factors** | | |
| Exposure to air pollution | Does the patient *feel they* are exposed to high levels of air or traffic pollution? | - |
| Exposure to hazardous substances or radiation | Is the patient exposed to toxic substances or chemicals in their home, workplace or surrounding environment *(such as pesticides, solvents, heavy metals, tobacco smoke, carbon monoxide, ionising radiation)*? | “It is not clear to me what toxic substances are, what that could be. I think that could be really hard for people to answer, cause some people wouldn't really know what they were. Maybe some examples can be included related to home and so on.” (PRA) |

PRA: quote from person of reproductive age; PCP: quote from primary care professional.

Text in *italics* are changes made to preconception health topics and example questions / prompts.

**Supplementary Table 7.** Characteristics of Delphi survey participants at each round

|  | **Delphi survey round 1**  **N = 85** | | **Delphi survey round 2**  **N = 69** | | **Delphi survey round 3**  **N = 63** | |
| --- | --- | --- | --- | --- | --- | --- |
|  | **People of reproductive age**  **n = 48** | **Primary care practitioners**  **n = 37** | **People of reproductive age**  **n = 39** | **Primary care practitioners**  **n = 30** | **People of reproductive age**  **n = 35** | **Primary care practitioners**  **n = 28** |
| **Place of residence, n (%)** |  |  |  |  |  |  |
| England | 33 (68.6) | 19 (50.0) | 28 (71.8) | 16 (53.3) | 26 (74.3) | 15 (53.6) |
| Wales | 3 (5.7) | 4 (10.7) | 2 (5.1) | 3 (10.0) | 2 (5.7) | 3 (10.7) |
| Scotland | 5 (11.4) | 7 (17.9) | 4 (10.3) | 5 (16.7) | 3 (8.6) | 5 (17.9) |
| Northern Ireland | 7 (14.3) | 8 (21.4) | 5 (12.8) | 6 (20.0) | 4 (11.4) | 5 (17.9) |
| **Age group, n (%)** |  |  |  |  |  |  |
| 16-20 years | 3 (5.7) | - | 2 (5.1) | - | 2 (5.7) | - |
| 21-30 years | 11 (22.9) | - | 9 (23.1) | - | 9 (25.7) | - |
| 31-40 years | 18 (37.1) | 19 (50.0) | 14 (35.9) | 17 (56.7) | 13 (37.1) | 15 (53.6) |
| 41-50 years | 16 (34.3) | 9 (25.0) | 14 (35.9) | 7 (23.3) | 11 (31.4) | 6 (21.4) |
| 51-60 years | *NA* | 5 (14.3) | *NA* | 4 (13.3) | *NA* | 4 (14.3) |
| 61-70 years | *NA* | 1 (3.6) | *NA* | 1 (3.3) | *NA* | 1 (3.6) |
| Over 70 years | *NA* | 1 (3.6) | *NA* | 1 (3.3) | *NA* | 1 (3.6) |
| Prefer not to answer | - | 1 (3.6) | - | - | - | - |
| **Gender, n (%)** |  |  |  |  |  |  |
| Woman | 29 (60.0) | 24 (64.3) | 25 (64.1) | 20 (66.7) | 24 (68.6) | 20 (71.4) |
| Man | 14 (28.6) | 13 (35.7) | 10 (25.6) | 10 (33.3) | 8 (22.9) | 8 (28.6) |
| Non-binary | 3 (5.7) | - | 2 (5.1) | - | 1 (2.9) | - |
| Transgender | 3 (5.7) | - | 2 (5.1) | - | 2 (5.7) | - |
| **Ethnicity, n (%)** |  |  |  |  |  |  |
| White | 4 (8.6) | 4 (10.7) | 2 (5.1) | 2 (6.7) | 2 (5.7) | 2 (7.1) |
| White British | 21 (42.9) | 15 (39.3) | 18 (46.2) | 12 (40.0) | 16 (45.7) | 10 (35.7) |
| Asian | 7 (14.3) | 5 (14.3) | 5 (12.8) | 5 (16.7) | 4 (11.4) | 5 (17.9) |
| Asian British | 4 (8.6) | 5 (14.3) | 4 (10.3) | 4 (13.3) | 4 (11.4) | 4 (14.3) |
| Black | 3 (5.7) | 3 (7.1) | 3 (7.7) | 2 (6.7) | 3 (8.6) | 2 (7.1) |
| Black British | 3 (5.7) | 1 (3.6) | 2 (5.1) | 1 (3.3) | 2 (5.7) | 1 (3.6) |
| Mixed | 5 (11.4) | 4 (10.7) | 4 (10.3) | 4 (13.3) | 3 (8.6) | 1 (3.6) |
| Other: Arab | 1 (2.9) | - | 1 (2.6) | - | 1 (2.9) | 4 (14.3) |
| **Previously been pregnant or currently pregnant, n (%)** | 23 (47.9) | *NA* | 19 (48.7) | *NA* | 17 (48.6) | *NA* |
| **Pregnancy and parenthood intentions, n (%)** |  |  |  |  |  |  |
| Currently trying for a baby | 5 (10.4) | *NA* | 5 (12.8) | *NA* | 5 (14.3) | *NA* |
| Would like to try for a baby in the next year or so | 3 (6.3) | *NA* | 3 (7.7) | *NA* | 3 (8.6) | *NA* |
| Would like to try for a baby, but not yet in the next year | 7 (14.6) | *NA* | 6 (15.4) | *NA* | 5 (14.3) | *NA* |
| Would like to try, but struggling to get pregnant | 5 (10.4) | *NA* | 5 (12.8) | *NA* | 5 (14.3) | *NA* |
| Not sure | 7 (14.6) | *NA* | 5 (12.8) | *NA* | 3 (8.6) | *NA* |
| Already have children, don’t want to have more | 15 (33.3) | *NA* | 10 (25.6) | *NA* | 9 (25.7) | *NA* |
| Don’t have children, and do not want them | 3 (6.3) | *NA* | 3 (7.7) | *NA* | 3 (8.6) | *NA* |
| Other: already have a child and unable to have more | 1 (2.1) | *NA* | 1 (2.6) | *NA* | 1 (2.9) | *NA* |
| Other: hoping to adopt | 1 (2.1) | *NA* | 1 (2.6) | *NA* | 1 (2.9) | *NA* |
| **Long-term health condition, n (%)** |  |  |  |  |  |  |
| Physical health condition | 10 (20.0) | *NA* | 8 (20.5) | *NA* | 6 (17.1) | *NA* |
| Mental health condition | 7 (14.3) | *NA* | 6 (15.4) | *NA* | 6 (17.1) | *NA* |
| Physical and mental health condition | 7 (14.3) | *NA* | 5 (12.8) | *NA* | 5 (14.3) | *NA* |
| No long-term health condition | 23 (48.6) | *NA* | 20 (51.3) | *NA* | 18 (51.4) | *NA* |
| Prefer not to answer | 1 (2.9) | *NA* | - | *NA* | - | *NA* |
| **General practice contact, n (%)** |  |  |  |  |  |  |
| At least once every year | 37 (77.1) | *NA* | 33 (84.6) | *NA* | 31 (88.6) | *NA* |
| Most years, but not every year | 11 (22.9) | *NA* | 6 (15.4) | *NA* | 4 (11.4) | *NA* |
| **Main profession, n (%)** |  |  |  |  |  |  |
| GP | *NA* | 17 (46.4) | *NA* | 13 (43.3) | *NA* | 11 (39.3) |
| Nurse | *NA* | 4 (10.7) | *NA* | 4 (13.3) | *NA* | 4 (14.3) |
| Pharmacist | *NA* | 3 (7.1) | *NA* | 2 (6.7) | *NA* | 2 (7.1) |
| Midwife | *NA* | 1 (3.6) | *NA* | 1 (3.3) | *NA* | 1 (3.6) |
| Health visitor | *NA* | 3 (7.1) | *NA* | 2 (6.7) | *NA* | 2 (7.1) |
| Link worker / social prescriber | *NA* | 3 (7.1) | *NA* | 2 (6.7) | *NA* | 2 (7.1) |
| Other: practice manager | *NA* | 3 (7.1) | *NA* | 2 (6.7) | *NA* | 2 (7.1) |
| Other: GP trainee / junior doctor / GP registrar | *NA* | 3 (7.1) | *NA* | 3 (10.0) | *NA* | 3 (10.7) |
| Other: Public Health Officer | *NA* | 1 (3.6) | *NA* | 1 (3.3) | *NA* | 1 (3.6) |
| **Professional experience, n (%)** |  |  |  |  |  |  |
| Less than 5 years | *NA* | 15 (39.3) | *NA* | 12 (40.0) | *NA* | 11 (39.3) |
| 5-10 years | *NA* | 7 (17.9) | *NA* | 6 (20.0) | *NA* | 5 (17.9) |
| 10-15 years | *NA* | 4 (10.7) | *NA* | 3 (10.0) | *NA* | 3 (10.7) |
| 15+ years | *NA* | 12 (32.1) | *NA* | 9 (30.0) | *NA* | 9 (32.1) |

*NA*, not applicable, i.e. question not asked.

**Supplementary Table 8**. Delphi survey results: proportion of participants scoring topics as ‘no or limited importance’ (score 1-3) or ‘critically important’ (score 7-9) across all three survey rounds.

|  | **Delphi survey round 1** | | | | **Delphi survey round 2** | | | | **Delphi survey round 3** | | | |
| --- | --- | --- | --- | --- | --- | --- | --- | --- | --- | --- | --- | --- |
|  | **People of reproductive age** | | **Primary care practitioners** | | **People of reproductive age** | | **Primary care practitioners** | | **People of reproductive age** | | **Primary care practitioners** | |
|  | **Score 1-3 (%)** | **Score 7-9 (%)** | **Score 1-3 (%)** | **Score 7-9 (%)** | **Score 1-3 (%)** | **Score 7-9 (%)** | **Score 1-3 (%)** | **Score 7-9 (%)** | **Score 1-3 (%)** | **Score 7-9 (%)** | **Score 1-3 (%)** | **Score 7-9 (%)** |
| Pregnancy intention and timing | 13 | 35 | 0 | 67 | 18 | 41 | 0 | 29 | 14 | 55 | 8 | 0 |
| Contraception use | 9 | 39 | 0 | 38 | 0 | 35 | 0 | 43 | 0 | 46 | 14 | 0 |
| Pregnancy anxiety | 4 | 43 | 8 | 31 | 6 | 47 | 0 | 29 | 11 | 49 | 21 | 0 |
| Infertility issues and treatment | 4 | **78** | 0 | 31 | 0 | **71** | 0 | 29 | 0 | 62 | 0 | 0 |
| Previous pregnancy complications and experiences | 0 | **74** | 0 | **82** | 0 | **71** | 0 | **71** | NA | NA | NA | NA |
| Previous pregnancy or baby loss | 4 | **83** | 0 | **83** | 6 | **88** | 0 | **71** | NA | NA | NA | NA |
| Mental health | 0 | **79** | 0 | 67 | 0 | **83** | 0 | **71** | NA | NA | NA | NA |
| Physical health | 0 | **91** | 0 | **75** | 0 | **89** | 0 | **71** | NA | NA | NA | NA |
| Neurodiversity | 8 | 42 | 17 | 25 | 11 | 28 | 29 | 29 | 17 | 31 | 43 | 0 |
| Medication use | 0 | **79** | 0 | **92** | 0 | **89** | 0 | **100** | NA | NA | NA | NA |
| Gender identity and gender transition | 22 | 26 | 17 | 50 | 12 | 41 | 29 | 57 | 23 | 38 | 25 | 14 |
| Genetic conditions | 0 | **71** | 0 | 67 | 0 | **72** | 0 | **71** | NA | NA | NA | NA |
| Weight status | 8 | 46 | 0 | **92** | 6 | 44 | 0 | **100** | 0 | 69 | 0 | 43 |
| Previous bariatric surgery | 17 | 26 | 25 | 17 | 13 | 19 | 14 | 14 | 17 | 25 | 29 | 0 |
| Nutritional deficiencies | 0 | 54 | 0 | 50 | 0 | 53 | 0 | **71** | 0 | 69 | 0 | 14 |
| Dental health | 29 | 21 | 50 | 17 | 35 | 0 | 29 | 14 | 40 | 0 | 39 | 0 |
| Cervical smear status | 14 | 41 | 17 | 17 | 19 | 44 | 14 | 0 | 26 | 17 | 43 | 7 |
| Immunisation status | 17 | 38 | 8 | 42 | 18 | 35 | 0 | 57 | 0 | 46 | 0 | 29 |
| Infectious diseases | 0 | **71** | 0 | 69 | 0 | 59 | 0 | 57 | 0 | **77** | 0 | 14 |
| Sexually transmitted infections | 4 | 67 | 0 | **83** | 0 | **71** | 0 | **71** | NA | NA | NA | NA |
| Other health services | 5 | 48 | 23 | 23 | 7 | 29 | 0 | 43 | 8 | 42 | 14 | 0 |
| Dietary habits | 4 | 61 | 0 | 25 | 0 | 56 | 0 | 43 | 0 | 54 | 0 | 7 |
| Folic acid supplement use | 5 | 64 | 0 | **100** | 6 | 67 | 0 | **100** | 0 | **77** | 0 | 43 |
| Dietary supplement use | 13 | 48 | 23 | 8 | 11 | 33 | 0 | 14 | 14 | 23 | 29 | 0 |
| Physical activity level | 0 | 39 | 8 | 25 | 0 | 33 | 0 | 29 | 0 | 38 | 7 | 0 |
| Smoking status | 0 | **91** | 0 | **100** | 0 | **94** | 0 | **100** | NA | NA | NA | NA |
| Alcohol consumption | 0 | **96** | 0 | **100** | 0 | **89** | 0 | **100** | NA | NA | NA | NA |
| Drug use | 0 | **91** | 0 | **100** | 0 | **94** | 0 | **100** | NA | NA | NA | NA |
| Living circumstances and financial security | 13 | 39 | 31 | 15 | 12 | 35 | 14 | 14 | 17 | 23 | 14 | 14 |
| Physical, sexual and emotional abuse | 4 | **74** | 0 | **85** | 6 | **76** | 0 | **86** | NA | NA | NA | NA |
| Social support | 9 | 39 | 0 | 15 | 6 | 41 | 0 | 14 | 0 | 31 | 0 | 11 |
| Partner support and relationship | 13 | 43 | 8 | 17 | 6 | 41 | 0 | 29 | 23 | 38 | 0 | 0 |
| Access to care | 4 | 65 | 0 | 45 | 0 | **71** | 0 | 17 | 17 | 49 | 0 | 11 |
| Knowledge of preconception health and pregnancy | 9 | 52 | 0 | 38 | 12 | 41 | 0 | 43 | 6 | 54 | 0 | 0 |
| Ethnicity | 35 | 9 | 23 | 31 | 35 | 6 | 43 | 29 | 31 | 8 | 57 | 0 |
| Exposure to air pollution | 26 | 22 | 31 | 8 | 44 | 11 | 29 | 14 | 31 | 8 | 71 | 0 |
| Exposure to dangerous substances or radiation | 13 | 61 | 8 | 69 | 28 | 56 | 14 | 57 | 23 | 54 | 29 | 29 |

Score 1-3: ‘no or limited importance’

Score 7-9: ‘critically important’

Bolded numbers in grey boxes indicate ≥70% of participants scoring the topic as ‘critically important’

**Supplementary Table 9**. Voting results following final consensus workshop breakout session 1^a^

| **Topic** | **Votes for ‘No or limited importance’** | **Votes for ‘Important but not critical’** | **Votes for ‘Critically important’** |
| --- | --- | --- | --- |
| **Consensus in** |  |  |  |
| Weight status | 1 (4.8) | 4 (19.0) | **16 (76.2)** |
| Folic acid supplement use | 0 (0.0) | 0 (0.0) | **21 (100.0)** |
| Knowledge of preconception health and pregnancy | 0 (0.0) | 1 (4.8) | **20 (95.2)** |
| **Consensus out** |  |  |  |
| Exposure to air pollution | **18 (85.7)** | 3 (14.3) | 0 (0.0) |
| Exposure to dangerous substances or radiation | **16 (76.2)** | 5 (23.8) | 0 (0.0) |
| **No consensus** |  |  |  |
| Pregnancy intention and timing | 3 (14.3) | 11 (52.4) | 7 (33.3) |
| Contraception use | 10 (47.6) | 10 (47.6) | 1 (4.8) |
| Pregnancy anxiety | 0 (0.0) | 14 (66.7) | 7 (33.3) |
| Infertility issues and treatment | 1 (4.8) | 9 (42.9) | 11 (52.4) |
| Neurodiversity | 13 (61.9) | 8 (38.1) | 0 (0.0) |
| Gender identity and gender transition | 9 (42.9) | 12 (57.1) | 0 (0.0) |
| Previous bariatric surgery (weight loss surgery) | 8 (38.1) | 12 (57.1) | 1 (4.8) |
| Nutritional deficiencies | 2 (9.5) | 16 (76.2) | 3 (14.3) |
| Dental health | 14 (66.7) | 6 (28.6) | 1 (4.8) |
| Cervical smear status | 13 (61.9) | 8 (38.1) | 0 (0.0) |
| Immunisation status | 5 (23.8) | 6 (28.6) | 10 (47.6) |
| Infectious diseases | 1 (4.8) | 20 (95.2) | 0 (0.0 |
| Known to other health services | 7 (33.3) | 13 (61.9) | 1 (4.8) |
| Dietary habits | 9 (42.9) | 9 (42.9) | 3 (14.2) |
| Dietary supplement use | 5 (23.8) | 14 (66.7) | 2 (9.5) |
| Physical activity level | 1 (4.8) | 14 (66.7) | 6 (28.6) |
| Living circumstances and financial security | 7 (33.3) | 12 (57.1) | 2 (9.5) |
| Social support | 4 (19.0) | 16 (76.2) | 1 (4.8) |
| Partner support and relationship | 3 (14.3) | 14 (66.7) | 4 (19.0) |
| Access to care | 4 (19.0) | 15 (71.4) | 2 (9.5) |
| Ethnicity | 7 (33.3) | 11 (52.4) | 3 (14.3) |

^a^ Bolded numbers indicate ≥70% of participants scoring the topic as ‘critically important’

**Supplementary Table 10**. Voting results following final consensus workshop breakout session 2^a^

| **Topic** | **Votes for ‘Yes – a priority topic’** | **Votes for ‘No – not a priority topic’** |
| --- | --- | --- |
| **Consensus in** |  |  |
| Pregnancy intention and timing | **15 (71.4)** | 6 (25.6) |
| Immunisation status | **16 (76.2)** | 5 (23.8) |
| **Consensus out** |  |  |
| Pregnancy anxiety | 4 (19.0) | **17 (81.0)** |
| Neurodiversity | 1 (4.8) | **20 (95.2)** |
| Gender identity and gender transition | 1 (4.8) | **20 (95.2)** |
| Previous bariatric surgery (weight loss surgery) | 1 (4.8) | **20 (95.2)** |
| Nutritional deficiencies | 5 (23.8) | **16 (76.2)** |
| Dental health | 3 (14.3) | **18 (85.7)** |
| Cervical smear status | 2 (9.5) | **19 (90.5)** |
| Infectious diseases | 4 (19.0) | **17 (81.0)** |
| Known to other health services | 0 (0.0) | **21 (100.0)** |
| Dietary habits | 1 (4.8) | **20 (95.2)** |
| Dietary supplement use | 3 (14.3) | **18 (85.7)** |
| Physical activity level | 4 (19.0) | **17 (81.0)** |
| Living circumstances and financial security | 5 (23.8) | **16 (76.2)** |
| Social support | 4 (19.0) | **17 (81.0)** |
| Partner support and relationship | 5 (23.8) | **16 (76.2)** |
| Access to care | 4 (19.0) | **17 (81.0)** |
| Ethnicity | 5 (23.8) | **16 (76.2)** |
| **No consensus** |  |  |
| Contraception use | 7 (33.3) | 14 (66.7) |
| Infertility issues and treatment | 13 (61.9) | 8 (38.1) |

^a^ Bolded numbers indicate ≥70% of participants scoring the topic as ‘Yes – a priority topic’ or ‘No – not a priority topic’
